# Socioeconomic determinants of virtual care use among people living with HIV in a clinical cohort in Ontario, Canada: A cross-sectional study

**DOI:** 10.1101/2025.06.10.25329370

**Authors:** Nadia Rehman, Lawrence Mbuagbaw, Dominik Mertz, Giulia M. Muraca, Aaron Jones, the Ontario HIV Treatment Network Cohort Study

## Abstract

**Background:** Engagement in HIV care significantly improves health outcomes and life expectancy. However, socioeconomic challenges often impeder consistent care. Virtual care offers a potential solution by enhancing timely access to HIV services and addressing these barriers.

**Objectives:** We aimed to examine the association between socioeconomic factors and the use of virtual care among people living with HIV (PLHIV) in a clinical cohort in Ontario, Canada.

**Methods:** We analysed 2022 data from the Ontario HIV Treatment Network Cohort Study (OCS), coinciding with the initial rollout of virtual care. The OCS is a multi-site cohort comprising patients from 10 HIV clinics, with data obtained from medical charts, interviews, and linkage to provincial public health lab (PHOL) records for viral load testing. Multinomial logistic regression was used to identify predictors of care mode: virtual, in-person, or a combination of both.

**Results:** The study included 1,930 participants. Of these, 19% (n = 367) received virtual care, 45.6% (n = 900) received in-person care, and 34.3% (n = 663) accessed both virtual and in-person services. The median participant age was 55 years [Q1; Q3: 45; 62], and 78% (n = 1,131) identified as men who have sex with men (MSM).

Compared to Toronto residents, individuals living in Southwestern Ontario had higher odds of using virtual care (adjusted OR (AOR) 1.67, 95% CI: 1.13, 2.47). Conversely, females (AOR = 0.59; 95% CI: 0.40, 0.88), non-MSM men (AOR = 0.64; 95% CI: 0.45, 0.92), residents of Eastern Ontario (AOR = 0.42; 95% CI: 0.26, 0.68), individuals with a high school education compared to those with a university degree (AOR = 0.67; 95% CI: 0.46, 0.98), those with an annual gross income of CAD $71,000-90,000 (AOR = 0.59; 95% CI: 0.38, 0.91), and individuals diagnosed with HIV within the last 10 years compared to those diagnosed more than 10 years ago (AOR = 0.59; 95% CI: 0.39, 0.91) were less likely to use virtual care. Participants experiencing any level of depression were more likely to use both virtual and in-person care options.

**Conclusion:** Virtual care was introduced during the COVID-19 pandemic to enhance healthcare access in Ontario. Its adoption varied based on socioeconomic and health-related factors in the OCS cohort. Ongoing research is needed to assess these patterns beyond the pandemic context..

## Introduction

Retention in HIV care is essential for the effective management of people living with HIV (PLHIV). While antiretroviral therapy (ART) has significantly reduced HIV-related morbidity and mortality, inconsistent retention in care can lead to low adherence to ART, drug resistance, poor health outcomes, and a greater risk of HIV transmission. Ensuring continuous care is vital for optimizing treatment outcomes [1–3].

The HIV burden is disproportionately prevalent among specific key populations and racial groups [4, 5]. In Ontario, 2022 data show that 56.8% of new HIV cases occurred among gay, bisexual, and men who have sex with men (MSM), 29.8% among individuals of African, Caribbean, and Black descent, and 10.6% among people with a history of intravenous drug use [6] These populations often face various socioeconomic disparities and individual challenges, including substance use, mental health conditions, stigma, and discrimination, which contribute to suboptimal retention in healthcare services and adherence to ART [7–9]. Furthermore, structural barriers within the patient-provider relationship and clinic settings can disrupt the continuity of HIV care [10, 11]. According to the Centers for Disease Control and Prevention Report, 2020, retention in HIV care varied across 45 states in the USA, with the highest retention (87%) among gender identity minorities and the lowest (61.5%) among people who inject drugs (PWID) [12].

The COVID-19 pandemic accelerated the adoption of virtual care across Ontario’s healthcare systems [13]. Due to its flexibility, virtual care has the potential to improve access to HIV services by addressing socioeconomic barriers. Although concerns persist, such as connectivity issues and the inability to conduct objective tests, participants reported benefits including increased privacy, reduced transportation needs, and greater scheduling flexibility[14, 15]. While concerns persist regarding connectivity issues and limitations such as a lack of objective testing, participants highlighted advantages like increased privacy, reduced transportation needs, and greater control over scheduling [16, 17].

Our recent analysis of 2022 data from the Ontario HIV Treatment Network Cohort Study (OCS) found that virtual care was associated with improved adherence to ART and better viral suppression. However, the cross-sectional design limited our ability to infer causality or assess differences in impact across sociodemographic groups[18]. A retrospective study from the Johns Hopkins HIV Clinical Cohort (2010) compared clinic visit uptake during one year of in-person care with a year of exclusively virtual care during the pandemic. Although overall visit uptake declined with the virtual model, certain groups, such as older adults, men, Black patients, and individuals with a history of substance use disorder, showed improved engagement. Notably, in that study, neither patients nor providers could choose the mode of care delivery [19].

Given the relative novelty of virtual care, it is important to understand the factors influencing the type of care modality used, as this insight can help address barriers to care retention. This study aimed to examine whether various sociodemographic, structural and health-related factors were associated with the use of virtual care among PLHIV in the 2022 OCS cohort, when COVID-19 restrictions had been lifted and virtual care had been formally adopted as a standard care option in Ontario, Canada [20]. Since patient satisfaction is linked to improved quality of life (QoL) and subsequently enhanced important patient outcomes, we supplemented our analysis with patient satisfaction surveys integrated into the OCS questionnaire [21]. To ensure the study is grounded in the experiences of those it aims to serve, we partnered with Realize, a Canadian charity supporting people with HIV, to establish a community advisory board (CAB)[22]. By incorporating the voices and insights of PLHIV, we aim for findings that resonate with communities [23, 24].

## Methods

### Data Sources and Study Design

We conducted a cross-sectional study using data from participants in the OCS cohort in 2022. The OCS is a multisite clinical cohort that includes individuals receiving HIV care in Ontario, Canada. Detailed information about the cohort has been described elsewhere[25]. Demographic, clinical, and behavioural data were collected through annual interviews, while relevant clinical data were abstracted from medical records. We also conducted record linkage with the HIV viral load database at Public Health Ontario Laboratories (PHOL), the province’s sole provider of viral load testing. All participants provided written informed consent upon recruitment, and the cohort design and consent forms were approved by the Research Ethics Boards (REBs) of the University of Toronto and the participating study sites.

### Study Population

We used cohort data available as of 2022 to define our study population. Participants were eligible for this analysis if they were 16 years or older, had visited their HIV physician through any of the three care modalities—virtual, in-person visits, or a combination of both in 2022—and had completed the OCS questionnaire. Individuals with incomplete information regarding the type of care received were excluded from the analysis.

### Measures/Outcomes

HIV care was classified into three mutually exclusive categories: (i) in-person clinic visits; (ii) virtual care via telephone or video call; and (iii) a combination of both. Virtual care was defined as consultations with an HIV care physician by telephone or video call, as defined in the 2022 OCS Questionnaire.

Baseline demographic and clinical covariate data were extracted from the 2022 OCS questionnaire to examine potential predictors of retention in 2022. Age was analyzed both as a continuous and categorical variable, divided into four groups: ≤30, 31-40, 41-50, and >50 years. Sex and sexual orientation were combined into a single variable with three categories: females, male MSM, and male non-MSM [7]. Other covariates are defined as follows: race or ethnicity (African, Caribbean, or Black versus White versus others); language fluency, derived by combining two variables: Canadian-born and immigrants living ≥10 years, defined as participants with proficient English-speaking skills, versus immigrants living for < 10 years, defined as having below-average English-speaking skills; relationship status, categorized as stable (married, living common-law, or in a committed relationship) versus unstable (widowed, separated/divorced, or single); regions in Ontario categorized into four areas (Greater Toronto Area, Eastern Ontario, Northern Ontario, Southwestern Ontario); education level (elementary versus high school versus college versus higher education); employment status (employed versus unemployed); and annual personal income (≤$50,000, $51,000-$70,000, $71,000-$100,000, and >$100,000).

Barriers to virtual care were defined as privacy or lack of privacy, depending on the availability of a private space for attending the virtual visit. The housing situation was categorized as stable (condo, housing facility, shelter, or room in a motel or hotel) or unstable (correctional facility, couch surfing, or living on the street).

Clinical covariates extracted include adherence to ART (self-reported), categorized as optimal adherence (≥95%: never skipped or skipped more than three months ago) and suboptimal adherence (<95%, defined as missed doses within the past week, one to two weeks, two to four weeks ago, or one to three months ago)[26]; alcohol abuse, measured using the Alcohol Use Disorder Identification Test (AUDIT-10), with harmful alcohol use defined as a score of ≥8 regardless of gender/sex[27]; depression, measured by the Patient Health Questionnaire (PHQ) scale, with nine items, with depression defined as normal (0-2), mild (3-5), moderate (6-8), and severe depression (9-12) [28]; health-related QoL, assessed using the Short Form 36 Health Survey with two components: Mental Component Summary Score (MCS) and Physical Component Summary Score (PCS) [29]; smoking, classified as heavy smokers (at least 20 cigarettes daily) vs. moderate smokers [30]; diagnosis of mental health comorbidities, based on self-report in the OCS questionnaire; and stigma, assessed using a 10-item HIV-related stigma scale, categorized into four major components: personalized stigma, worries about disclosure of status, negative self-image, and sensitivity to public reactions regarding HIV status. Individuals who responded with “agree” or “strongly agree” were identified as experiencing stigma in at least one of the four components [31].

### Patient satisfaction survey

The OCS questionnaire includes a satisfaction survey on virtual and in-person HIV care, divided into three sections:

### Experience with Virtual Care

Assessed across 10 elements: (1) physician’s time; (2) time saved; (3) cost savings; (4) involvement in decision-making; (5) sense of safety; (6) ease of technology use; (7) addressing health concerns; (8) communication of health issues; (9) preference for future virtual care; and (10) opportunity to ask treatment-related questions. Responses were rated on a 5-point Likert scale: strongly agree, agree, neither agree nor disagree, disagree, and strongly disagree, with higher scores indicating greater satisfaction.

#### Experience with HIV Care Provider

Evaluated 13 elements, including (1) familiarity with patient history; (2) listening skills; (3) language proficiency; (4) clarity in explanations; (5) sensitivity to needs; (6) dignity in treatment; (7) ability to provide instructions; (8) responsiveness to questions; (9) concern for medication coverage; (10) involvement in treatment; (11) adequate time spent; (12) confidence in the provider; and (13) confidence in provider knowledge. Responses used a 3-point Likert scale: excellent/very good, good, and fair/poor.

#### Experience with Clinical Practice

Assessed on five elements: (1) ease of accessing the visit; (2) wait time; (3) level of privacy; (4) consistency of communication; (5) quality of staff interactions: and (6) overall experience in accessing the care. Ratings followed the same 3-point Likert scale.

The type of HIV care received was summarized using numbers and percentages.

### Data analysis

Descriptive statistics were used, including proportions for categorical variables and medians with IQR for continuous variables. The continuous covariates were compared using Kruskal-Wallis, and categorical variables were compared using chi-square tests.

We used a three-category multinomial logistic regression, with in-person care as the reference category, to identify independent correlates of virtual care use [32]. Given the limited prior research on predictors of virtual care use, we included all covariates in the model irrespective of statistical significance, thereby creating an explanatory model. The model results are reported as odds ratios (OR) with 95% confidence intervals (CI). Since the OCS questionnaire had data missing at random, we performed 10 imputations for each model and combined the results using Rubin’s rule[33]. A p-value of <0.05 or a 95% CI excluding the null value was considered statistically significant. Statistical analysis was performed using R software version 4.4.1.

## Results

In 2022, a total of 2,155 people completed the OCS questionnaire, with 1,930 participants reporting the type of care they received. Of these, 19% (n = 367) of HIV care visits were conducted virtually, 46% (n = 900) were in-person visits, and 34% (n = 663) were a combination of both in-person and virtual care. The median age of the participants was 55 years (IQR: 45-62). A notable 78% (n = 1,497) of participants were men, with the majority identifying as MSM, making up 51.7% (1,119/1,930) of the sample. Among the MSM participants, 63% (708/1,119) optimally adhered to ART, with a median age of 57 years (IQR: 47-63).

In terms of racial demographics, participants identifying as White represented the largest group at 61% (1,172/1,930). The usage of the three different care modalities varied across participant characteristics.

Significant differences were observed in the proportions of in-person, virtual, and combined care usage based on sex, gender, race, availability of a private space for virtual visits, housing situation, ART adherence, insurance status, stigma status, depression status, and viral load (Table 1).

**Table 1.**
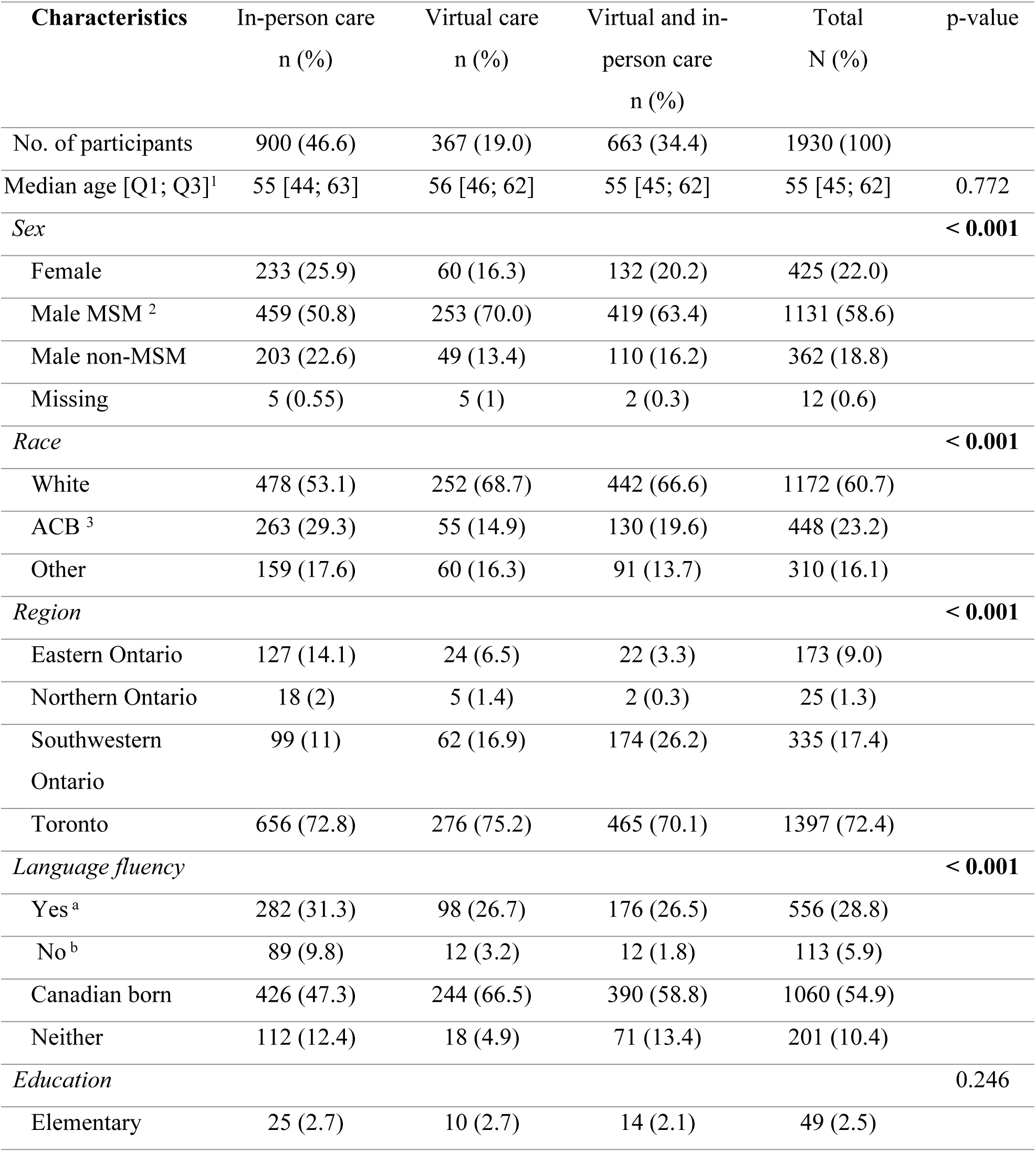
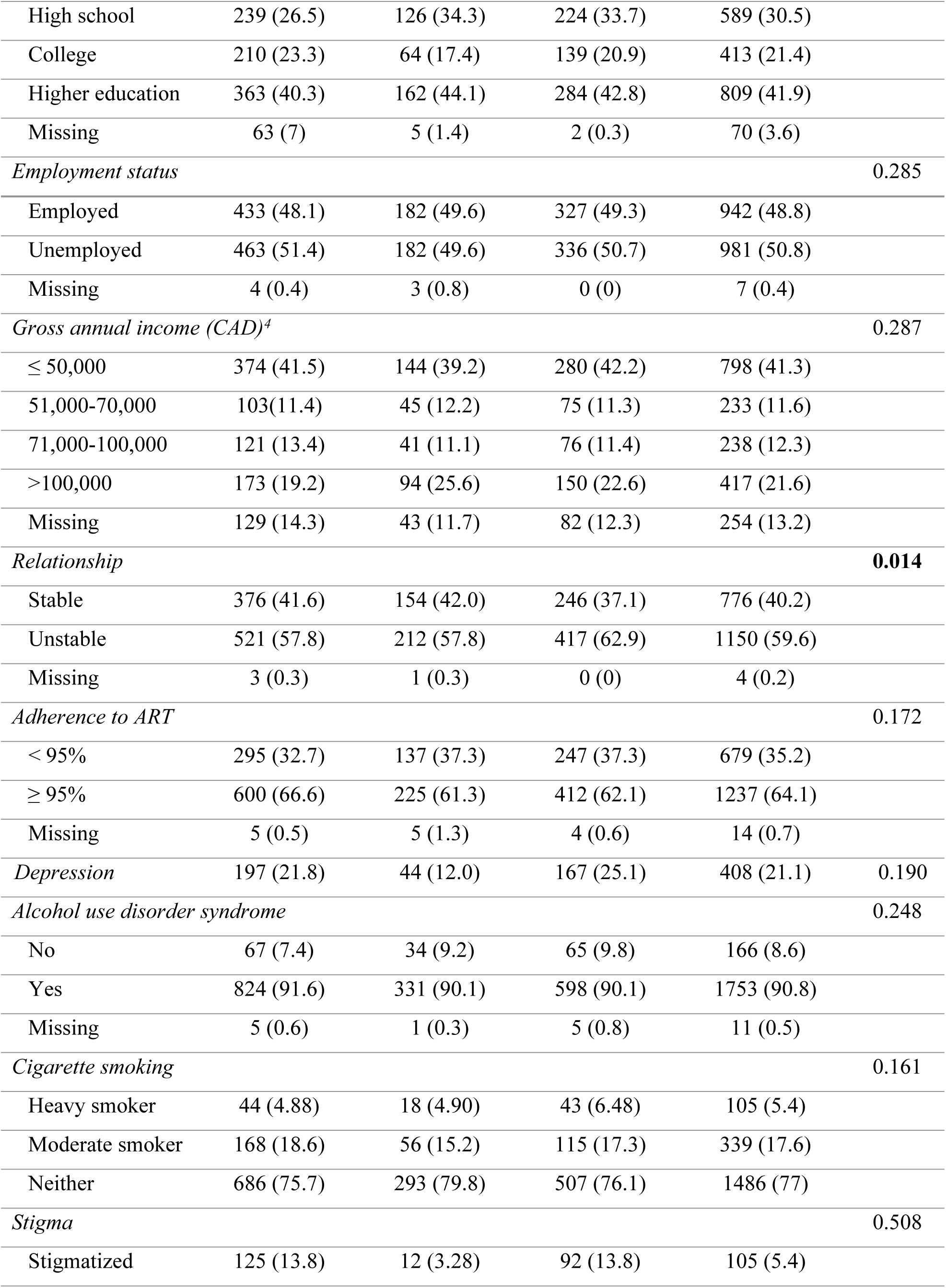
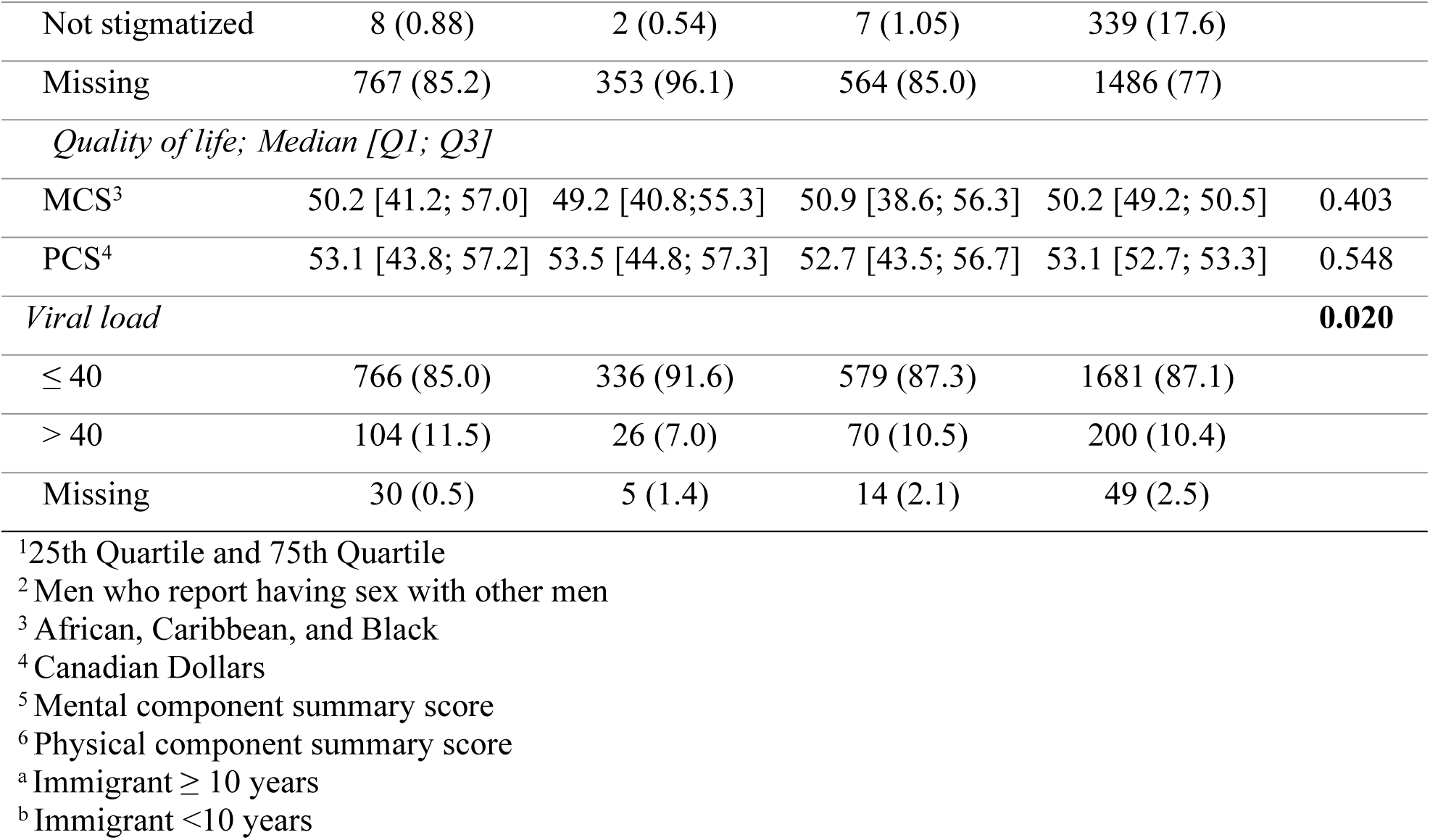
Comparison of baseline characteristics between participants who accessed HIV care through virtual, in-person or both virtual and in-person care in Ontario, Canada, in 2022

### Socio-demographic factors associated with the type of care

#### Sex

Compared to male MSM, female and male non-MSM participants were less likely to utilize virtual care than in-person care (females: Adjusted odds ratio (AOR) 0.59, 95% confidence interval (CI): 0.40, 0.88; and male non-MSM: AOR 0.64, 95% CI: 0.45, 0.92). Additionally, female and male non-MSM participants were less likely to use a combination of virtual and in-person visits compared to exclusively in-person visits (Females: AOR 0.65, 95% CI: 0.47, 0.90; male non-MSM: AOR 0.59, 95% CI: 0.40, 0.88).

#### Region

Compared to participants residing in Toronto, those in the Eastern region were less likely to receive virtual care (AOR 0.42, 95% CI: 0.26, 0.68) or both virtual and in-person care (AOR 0.23, 95% CI: 0.14, 0.37). In contrast, participants from the southwestern region of Ontario had higher odds of attending virtual care (AOR 1.67, 95% CI: 1.13, 2.47) or both virtual and in-person visits (AOR 1.67, 95% CI: 1.13, 2.47) compared to in-person care.

#### Education

Compared to individuals with a university degree, those with a high school education had lower odds of attending virtual visits (AOR 0.67, 95% CI: 0.46, 0.98). Participants with an annual gross income between $70,000 and $100,000 were less likely to attend virtual visits than those earning over $100,000 (AOR 0.59, 95% CI: 0.38, 0.91).

Participants diagnosed with HIV for less than 10 years were more likely to attend virtual visits compared to those diagnosed for more than 10 years. Additionally, participants with mild (AOR 2.20, 95% CI: 1.54, 3.13), moderate (AOR 2.06, 95% CI: 1.28, 3.31), moderate severe (AOR 2.80, 95% CI: 1.49, 5.26), and severe depression (AOR 2.46, 95% CI: 1.13, 5.32) all preferred a combination of virtual and in-person care rather than in-person care alone (Table 2).

**Table 2.**
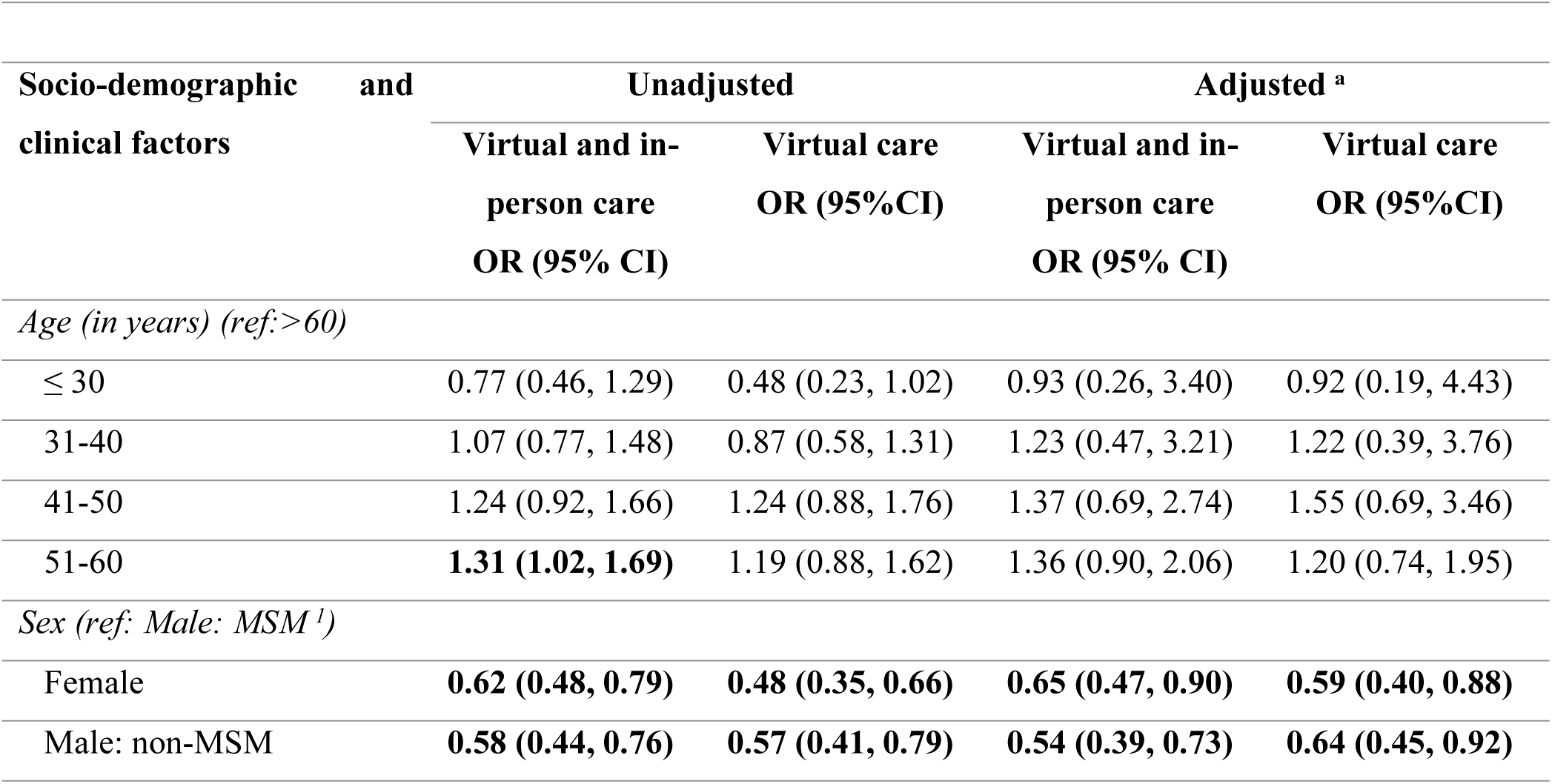
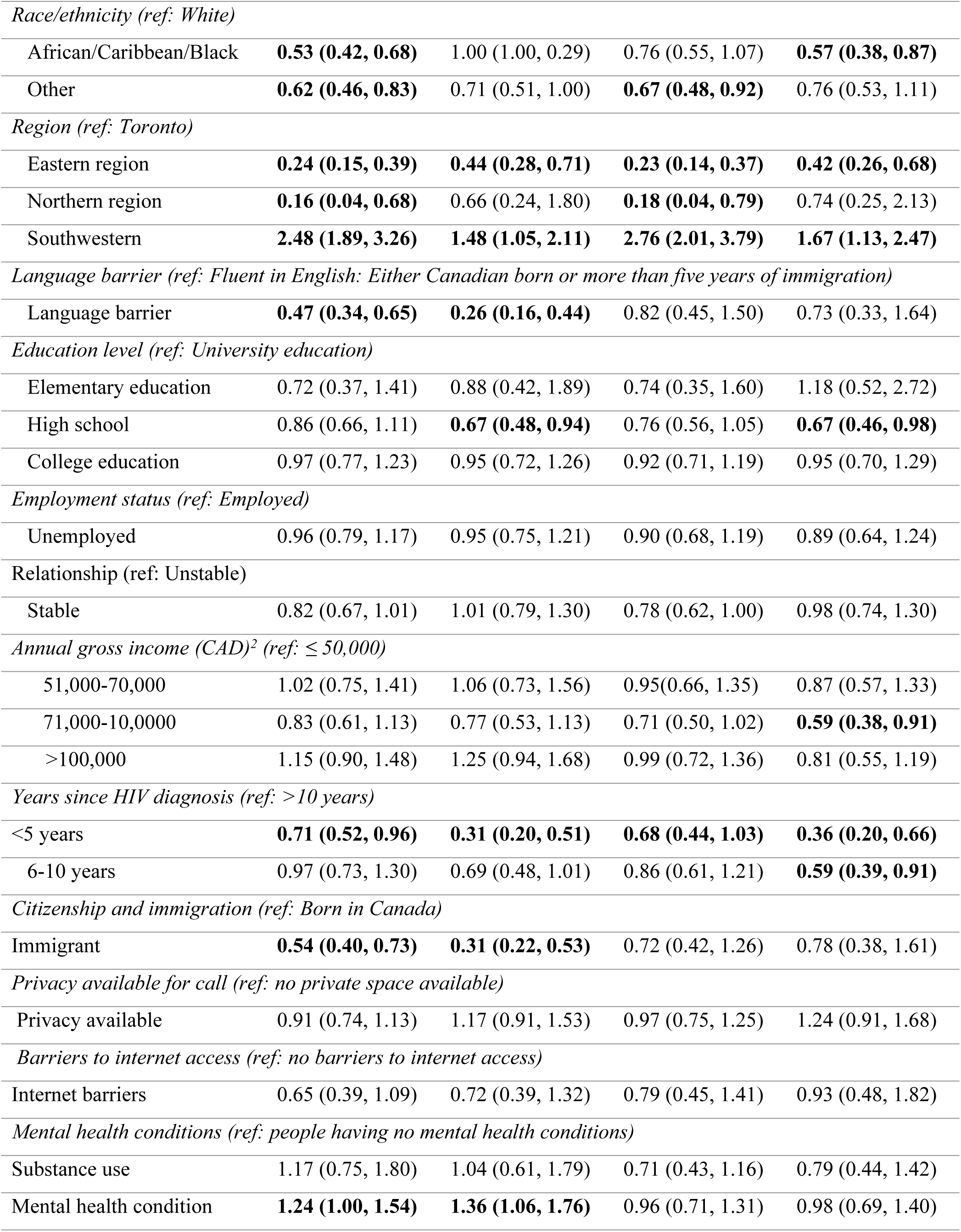
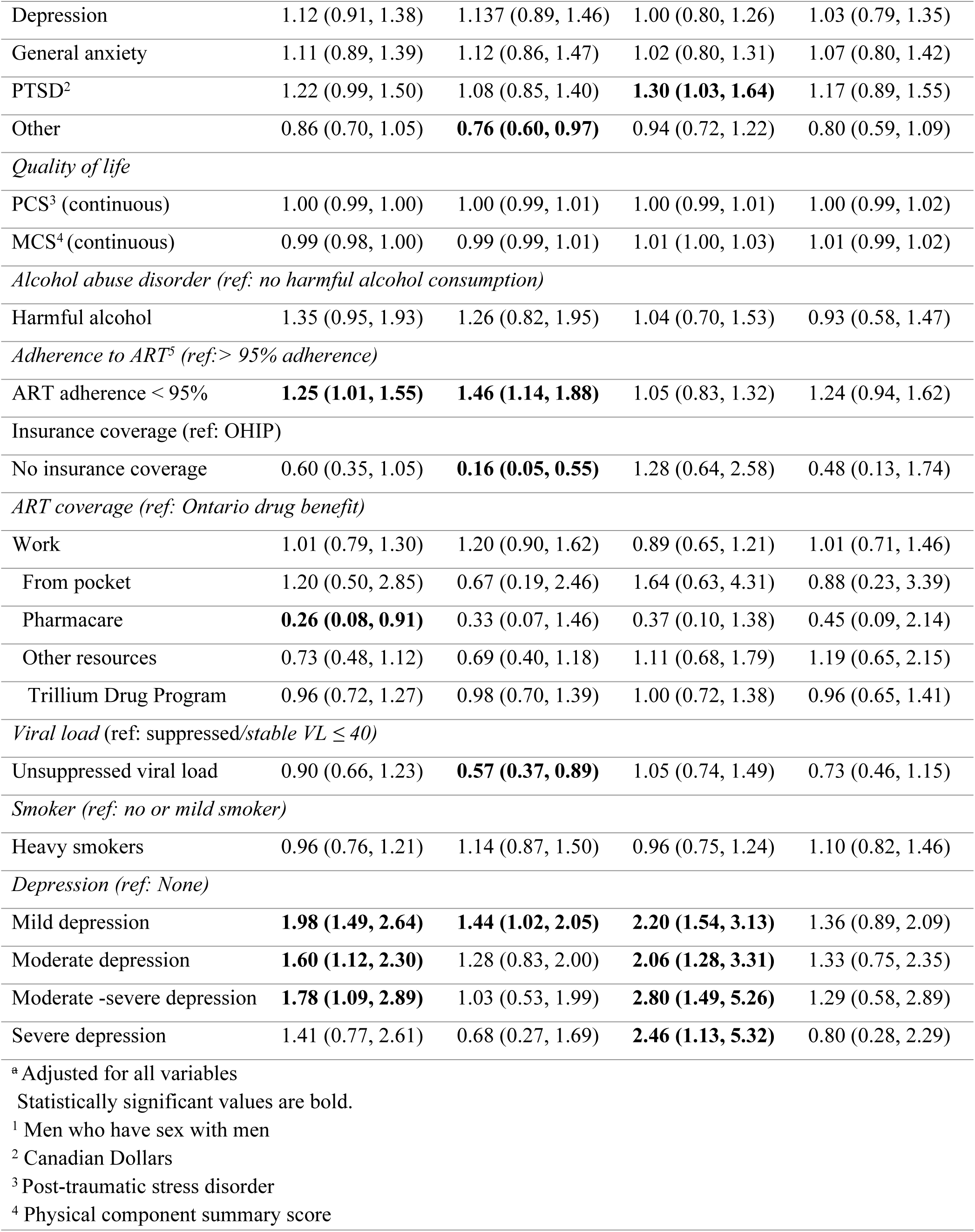
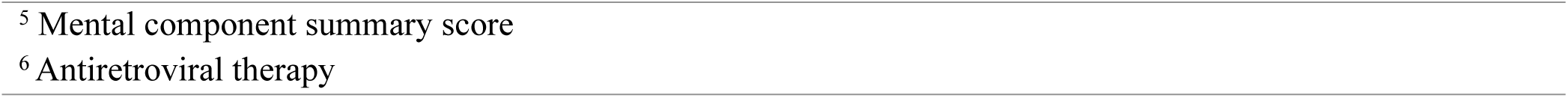
Factors associated with virtual care and virtual and in-person visits in HIV care (reference category: in-person visit) in 2022(n=1930)

#### Patients’ Satisfaction Survey

The survey on attitudes toward the virtual care experience revealed high satisfaction among participants using virtual care and both types of care, with 937 out of 1,030 participants (91.0%) reporting responses of “agree” or “strongly agree” across the 10 domains of the satisfaction survey (S1 Fig).

The survey on participants’ experiences with HIV clinical practices (S2 Fig) and care providers (S3 Fig) in all three groups provided high satisfaction; however, the response rate was only 22%.

The OCS questionnaire comprehensively covers various aspects of both patients’ and providers’ experiences. A statistically significant difference was observed in the number of participants who sought care from providers other than their usual HIV care provider. Among the groups, 41.2% of those using both virtual and in-person care (n = 454/630), 39.2% of those using in-person care (n = 600/900), and 54.9% of those using virtual care (n = 220/367) reported this (p-value < 0.05). The most common appointments were related to HIV care, including visits to family doctors, HIV specialists, pharmacists, and nurses.

#### Barriers to Virtual Visits

The most common reason participants opted for in-person visits was a preference for in-person consultations (n = 624/900, 69.3%), while 355/900 participants (40%) reported not being offered virtual care. Notably, 33.7% (652/1930) indicated they lacked a private space to attend virtual visits, with 17.1% (112/652) of those being part of the virtual care group. Further details regarding the factors influencing barriers to virtual visits are presented in Table 3.

**Table 3.**
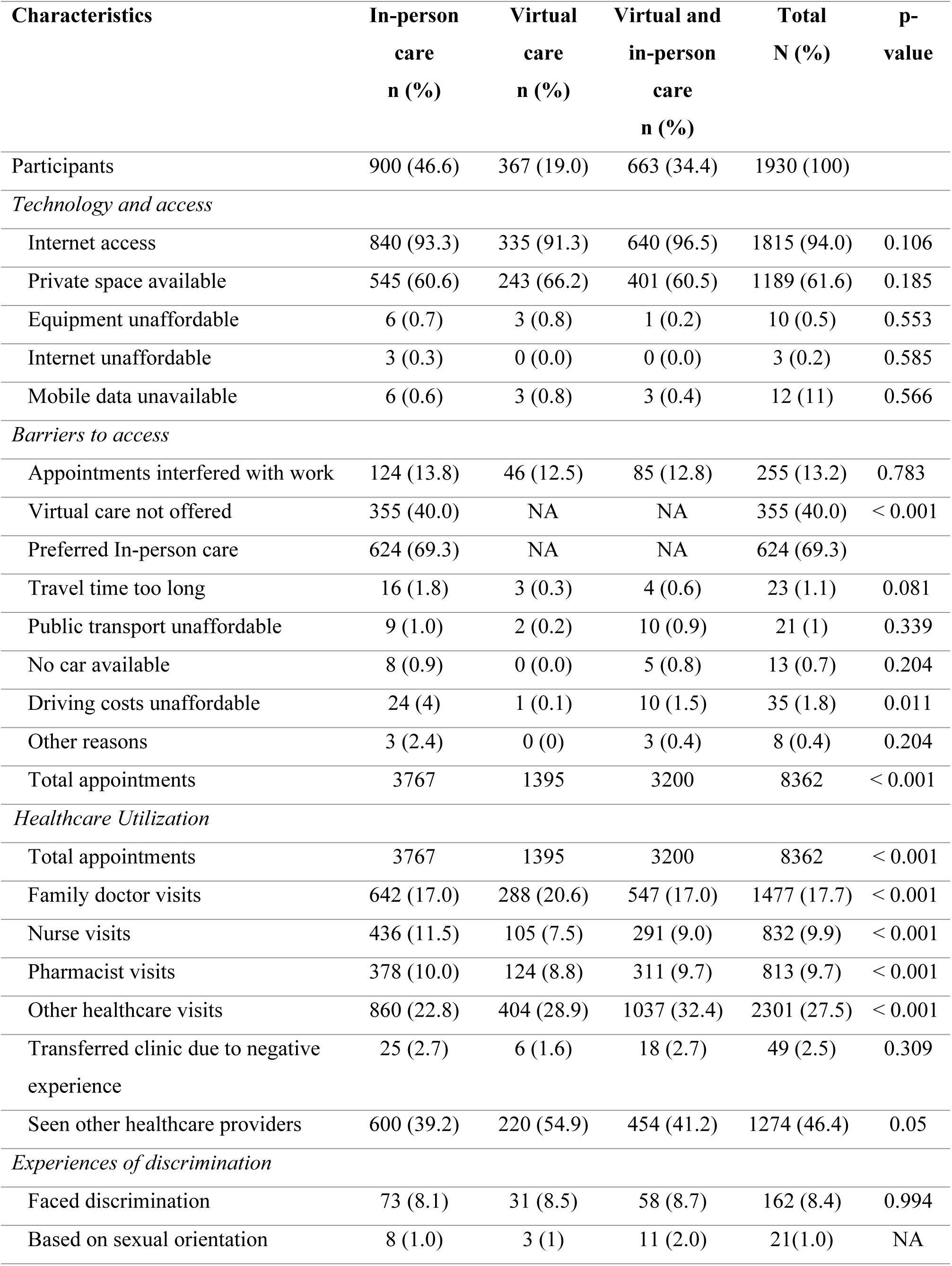
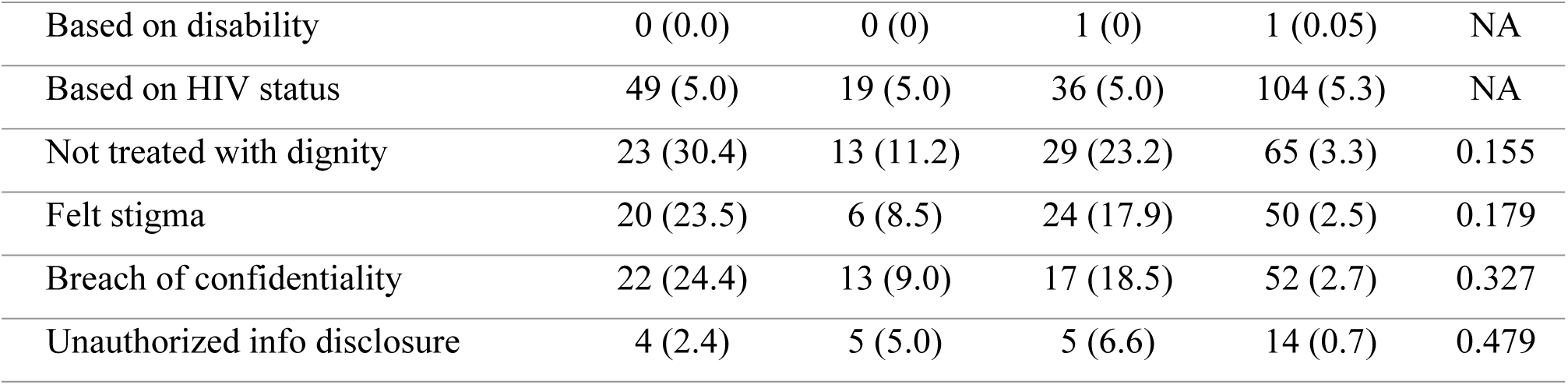
Factors influencing access to care by the mode of healthcare delivery.

## Discussion

This study examined three types of care in the 2022 OCS cohort: in-person, virtual, and a combination of both. The findings highlight a significant shift from in-person to virtual care, although in-person visits remained the predominant mode of care in the OCS cohort in 2022. Virtual care was accessed across all demographic and clinical groups, with higher usage observed among residents of Southwestern Ontario. In contrast, lower use of virtual care was associated with factors such as female sex, non-MSM status, residence in Eastern Ontario, a high school education, and an HIV diagnosis within the past 10 years. Patients with depression tended to use both virtual and in-person care, likely adjusting their care modality based on their mental health needs. However, the study could not determine whether the choice of care was linked to mental health status. Individuals aged 51-60 were the most frequent users of virtual care, suggesting greater technological comfort among older adults, although age-related differences were not statistically significant (p > 0.097). While previous research has linked employment to poor care retention, this study was conducted during the COVID-19 pandemic, a period when lockdowns likely influenced choices regarding care modalities. Further research is needed to assess the impact of virtual care on employment and access to care post-pandemic [34].

A study using data from the Veterans Health Administration examined clinical service use and patient characteristics during the pandemic. It found that individuals with lower income, greater disabilities, and more chronic conditions were more likely to use virtual care, while those experiencing homelessness were less likely to access it. In contrast, our study was conducted later, after restrictions had eased and virtual care had become more established. This provides a more accurate reflection of how HIV care modalities were chosen based on patient preferences, rather than external mandates or government-imposed restrictions.

To strengthen our assessment of virtual care, we incorporated a patient satisfaction survey within the OCS, focusing on key elements critical to continuity of care as defined by Shaw et al. [21]. Consistent with prior research [19]. Our findings indicate that satisfaction with virtual care is comparable to that with in-person care. Among virtual care users, 91.0% (937/1,028) reported satisfaction, indicating that it met their healthcare needs.

Regarding barriers to care, 70.4% (624/900) of in-person visits were based on personal preference, reflecting skepticism or limited trust in virtual healthcare. Additionally, 40% (355/900) reported not being offered virtual care, indicating a preference among providers for traditional consultations. Despite these barriers, satisfaction with HIV physicians and clinical settings remained consistently high across all groups.

### Limitations

This study has several limitations that affect the interpretation of its findings. Its cross-sectional design limits the ability to draw causal conclusions about the impact of virtual care on HIV outcomes. Furthermore, data were collected during the early stages of virtual care adoption, when standards, infrastructure, and provider training were still in development, which reduced the applicability of the results to current practices. To address these limitations, longitudinal research is needed, including cost analyses using linked administrative data to evaluate visit types and frequency over time [34, 35].

Although the patient satisfaction survey provides insights into care preferences, it primarily focuses on patient experience and access without capturing provider perspectives, which could introduce bias. Additionally, the study does not explore issues related to the usability and functionality of virtual care platforms. Future research should incorporate qualitative interviews and provider surveys to gain a deeper understanding of healthcare professionals’ experiences, preferences, and perceived limitations across various HIV care modalities.

### Strengths

Despite certain limitations, the study provides valuable insights into how sociodemographic and health factors influence HIV care choices. We examined the impact of various sociodemographic and health-related factors on access to care and explored the experiences, barriers, and acceptance of virtual care in detail. This offers a comprehensive overview and highlights avenues for future research and system improvements.

Although non-response bias is a common challenge in survey research, our study achieved a 22% response rate (427/1,930), which is sufficient to represent a population of 10,000 with a 5% margin of error. The use of a complete and accurate sampling frame, along with a validated survey instrument, further supports the reliability of the findings and enables population-level inferences while minimizing random sampling errors[36].

Drawing on a large sample and real-time data, the study highlights the importance of long-term research in fully assessing the potential of virtual care. While other studies have explored the experiences of PLHIV, highlighting the time- and cost-saving benefits and the ability to address disparities in care[37, 38] This is the first study to conduct a quantitative assessment of the virtual care use in relation to sociodemographic factors. Future studies should incorporate health administrative data to examine visit types, frequency, patient preferences, and physician perspectives regarding the care of people living with HIV over time.

## Conclusions

This study examined the use of virtual care among the 2022 OCS cohort of people living with HIV and the socio-demographic factors influencing their care preferences. Nearly half of the participants preferred traditional in-person visits, with male MSM, individuals with higher education and income, and those diagnosed with HIV for over 10 years showing a stronger preference. Regardless of the type of care utilized, patients reported high satisfaction with the services they received. Although confounded by COVID-19 lockdowns, this exploratory research paves the way for further investigation into the various modes of care delivery. Effective and equitable technology integration can enhance relationships, promote fair care, and improve outcomes for all PLHIV.

## Data Availability

The data is obtained from OHTN from the OCS study. It is unethical for the authors to share the data set publicly. However, the data request can be made directly with OHTN by submitting a Research Application Process (RAP) or please.

## Acknowledgements

The OHTN Cohort Study Team consists of Dr. Ann Burchell (Interim Principal Investigator), Unity Health and University of Toronto; Dr. Anita Benoit (Co-Investigator), University of Toronto; Dr. Lawrence Mbaugbaw (Co-Investigator), McMaster University; Dr. Sergio Rueda, CAMH and University of Toronto; Dr. Gordon Arbess, Unity Health; Dr. Corinna Quan, Windsor Regional Hospital; Dr. Curtis Cooper, Ottawa General Hospital; Elizabeth Lavoie and Dr. Maheen Saeed, Byward Family Health Team; Dr. Mona Loutfy and Dr. David Knox, Maple Leaf Medical Clinic; Dr. Nisha Andany, Sunnybrook Health Sciences Centre; Dr. Sharon Walmsley, University Health Network; Dr. Michael Silverman, St. Joseph’s Health Care; Tammy Bourque, Health Sciences North; Dr. Marek Smieja, Hamilton Health Sciences Centre; Wangari Tharao, Women’s Health in Women’s Hands Community Health Centre; Holly Gauvin, Elevate NWO; Dr. Jorge Martinez-Cajas, Kingston Hotel Dieu Hospital; and Dr. Jeffrey Craig, Lakeridge Positive Care Clinic.

We gratefully acknowledge all of the people living with HIV who volunteer to participate in the OHTN Cohort Study. We also acknowledge the work and support of OCS Governance Committee (Aaron Bowerman, Adrian Betts, Barry Adam, Cornel Gray, Dane Record, Jasmine Cotnam, Jason Brophy, Mary Ndung’u, Rodney Rousseau, Ruth Cameron, YY Chen) OCS Scientific Steering Committee (Anita Benoit, Ann Burchell, Barry Adam, Curtis Cooper, David Brennan, Kelly O’Brien, Lance Mcready, Lawrence Mbuagbaw, Mona Loutfy, Pierre Giguere, Sean Hillier, Sergio Rueda (Chair), and Trevor Hart) and Indigenous Data Governance Circle (Meghan Young, Randy Jackson, Trevor Stratton). The OHTN also acknowledges the work of past Governance Committee and Scientific Steering Committee members.

We thank all interviewers, data collectors, research associates, coordinators, nurses, and physicians who provide data collection support. The authors also wish to thank OCS staff for data management, IT support, and study coordination: Lucia Light, Mustafa Karacam, Nahid Qureshi, and Tsegaye Bekele. The Ontario Ministry of Health supports the OHTN Cohort Study.

We also acknowledge the Public Health Laboratories, Public Health Ontario, for supporting record linkage with the HIV viral load database.

The OHTN Cohort Study is supported by the Ontario Ministry of Health and Long-Term Care.

The opinions, results and conclusions are those of the authors and no endorsement by the Ontario HIV Treatment Network or Public Health Ontario is intended or should be inferred.

Realize provided support and guidance for CAB recruitment, consultation on community engagement and study design.

## Supporting information

**S1 Fig:** Bar graph on participants’ satisfaction with virtual care in the 2022 Ontario HIV Treatment Cohort Study (n = 1,030)

**S2 Fig** Bar graph on participants’ experiences with HIV care providers in the 2022 Ontario HIV Treatment Cohort Study (n = 432)

**S3 Fig:** Bar graph on participants’ HIV visits experiences with the clinical practice in the 2022 Ontario Treatment HIV Cohort Study (n = 427)

